# Neonatal Autonomic Regulation as a Predictor of Autism Spectrum Disorder in Very Preterm Infants

**DOI:** 10.1101/2023.11.14.23298262

**Authors:** Jessica Bradshaw, Christian O’Reilly, Kayla C. Everhart, Elizabeth Dixon, Amy Vinyard, Abbas Tavakoli, Victor Iskersky, Robin B. Dail

**Affiliations:** Department of Psychology, University of South Carolina, Columbia SC; Carolina Autism and Neurodevelopment Research Center, University of South Carolina, Columbia, SC; Department of Computer Science and Engineering, University of South Carolina, Columbia, SC; Artificial Intelligence Institute, University of South Carolina; College of Nursing, University of South Carolina, Columbia, SC; Prisma Health

## Abstract

Infants born preterm are at a significantly higher likelihood of having autism spectrum disorder (ASD). Preterm birth and ASD are both associated with neurological differences, notably autonomic nervous system (ANS) dysfunction, pointing to preterm ANS dysfunction as a potential pathway to ASD, particularly in VPT infants. In this study, a subset of very preterm (VPT) infants enrolled in a large, multisite clinical trial were enrolled in this study at birth (N=20). Continuous measures of minute-by-minute thermal gradients, defined by the difference between central and peripheral temperatures, and hour-by-hour abnormal heart rate characteristics (HRCs) were collected from birth-28 days (>40,000 samples/infant). Following NICU discharge, standardized measures of cognition, language, and motor skills were collected at adjusted ages 6, 9, and 12 months. At 12 months, assessments of social communication and early ASD symptoms were administered. Results suggest significant ASD concerns for half of the sample by 12 months of age. Neonatal abnormal HRCs were strongly associated with 12-month ASD symptoms (r=0.81, p<.01), as was birth gestational age (GA), birth weight (BW), and abnormal negative thermal gradients. ANS measures collected in the first month of neonatal life, more than a year prior to the ASD evaluation, were surprisingly strong predictors of ASD. This study highlights complementary ANS measures that describe how ANS dysfunction, likely resulting from an imbalance between the parasympathetic and sympathetic systems, may impact very early regulatory processes for neonates who later develop ASD. This finding offers a promising avenue for researching ANS-related etiological mechanisms and biomarkers of ASD.

## INTRODUCTION

Infants born preterm are at a significantly higher likelihood of having autism spectrum disorder (ASD), with reports of a 10-fold increase in the rate of ASD for very preterm infants (VPT, born < 32 weeks gestation)^1^ and a 20-fold increase for extremely preterm infants (born <28 weeks gestation)^2^. Yet, the etiological link between preterm birth and ASD remains unknown. Preterm birth and ASD are both associated with neurological differences, notably autonomic nervous system (ANS) dysfunction, pointing to neonatal ANS dysfunction as a promising early biomarker and potential pathway to ASD, particularly in VPT infants. The current study describes preliminary findings from a prospective, longitudinal trial of VPT infants with continuous measures of autonomic dysregulation in the first month of life, comprehensive neurodevelopmental monitoring, and ASD follow-up at age 1 year.

## METHODS

A subset of VPT infants enrolled in a large, multisite clinical trial^3^ were enrolled in this study at birth (N=20). Continuous measures of minute-by-minute thermal gradients, defined by the difference between central and peripheral temperatures^3^, and hour-by-hour abnormal heart rate characteristics (HRCs), measured using HeRO scores^4^, were collected from birth-28 days (>40,000 samples/infant). See additional details in Supplemental Information (SI). ANS dysfunction is indicated by abnormal HRCs (HeRO scores > 1) or elevated negative thermal gradients (peripheral > central body temperature)^5^. Following NICU discharge, standardized measures of cognition, language, and motor skills were collected at adjusted ages 6, 9, and 12 months (see SI). At 12 months, assessments of social communication and early ASD symptoms were administered^6–8^ (see SI). Study procedures were approved by hospital and university institutional review boards. Univariate, multivariate, and robust regression models were used to evaluate neonatal predictors of ASD outcomes.

## RESULTS

Descriptive statistics for all measures are presented in Tables 1 and S1. Continuous measures of ANS function were collected for N=20 infants enrolled at birth. Following NICU discharge, n=12 completed 1-year developmental follow-up (10 males) and n=1 was excluded from analyses due to significant medical morbidities that precluded a valid ASD assessment (see Fig. S1). Across all infants in the first 28 days of life, the average HeRO score was just above 1 and infants experienced abnormal negative thermal gradients for about 20% of the time (6 of 28 days). In terms of developmental trajectories, significant delays did not emerge until 12 months when over 50% of the sample exhibited social-communication delays and exceeded the clinical cutoff for ASD risk (see Table S1 and Fig S2). Neonatal abnormal HRCs were strongly associated with ASD symptoms at 12 months (*r*=0.81, *p*<.01; Fig. 1), as was birth gestational age (GA), birth weight (BW), and abnormal negative thermal gradients. A regression model including GA, BW, and abnormal HRCs revealed a significant, unique predictive effect of abnormal HRCs on ASD outcomes (model *r*=0.86, *p=*.017; HRC *r*=.72, *p*=.029).

**Table 1.** Descriptive Statistics and Correlations Between Neonatal Measures and ASD Symptoms.

|  | Mean (SD) | % Atypical<sup>a</sup> | Association with<br>ASD Outcome (Pearson's *r*)<sup>b</sup> | *p*-value |
| --- | --- | --- | --- | --- |
| *Neonatal Measures* |  |  |  |  |
| **Gestational Age (weeks)** | **28.25 (2.14)** | - | **-0.689** | **< .0001** |
| **Birthweight (g)** | **943.3 (227.49)** | - | **-0.499** | **.0031** |
| Medical Comorbidity Score<sup>c</sup> | 1.08 (0.99) | - | 0.241 | .474 |
| **Abnormal Heart Rate Characteristics (HeRO score)<sup>d</sup>** | **1.22 (0.63)** | - | **0.812** | **.002** |
| **Abnormal Negative Thermal Gradients (days)<sup>e</sup>** | **5.83 (1.18)** | - | **0.099** | **.039<sup>f</sup>** |
| *1-Year Developmental Outcomes* |  |  |  |  |
| Cognitive | 48.75 (11.39) | 8% | -0.332 | .318 |
| Gross Motor | 40.75 (11.99) | 50% | 0.028 | .935 |
| Receptive Language | 34.82 (7.76) | 64% | -0.385 | .243 |
| Expressive Language | 36.5 (12.46) | 58% | -0.170 | .183 |
| **Social Communication** | **74.27 (10.12)** | **72%** | **-0.732** | **.016** |
| *1-Year ASD Outcomes* |  |  |  |  |
| Parent-Reported ASD Symptoms | 16.95 (7.46) | 50% | - | - |
| ASD Symptom Composite | 19.00 (8.44) | 45% | - | - |
| ASD Red Flags | 7.18 (3.09) | 64% | - | - |

<sup>a</sup> % Atypical is defined as the percent of infants scoring below average on developmental outcomes and scoring in the “at-risk” range on the ASD measures.
<sup>b</sup> The primary ASD outcome in regression models is the Systematic Observation of ASD Red Flags (SORF)<sup>8</sup> Composite Score. This score is based on DSM-5 criteria for ASD, comprising both social interaction deficits and restricted interests and repetitive behaviors.
<sup>c</sup> Data on medical comorbidities was abstracted from the medical record and consists of the summative total of medical conditions experienced in the NICU (see SI).
<sup>d</sup> Abnormal heart rate characteristics are measured using HeRO scores. HeRO scores are calculated with a mathematical algorithm that quantifies abnormality using three components of heart activity: standard deviation of inter-heartbeat (RR) intervals; sample asymmetry; sample entropy.
<sup>e</sup> Number of days during which negative thermal gradients were present $\geq 30\%$ of the day.
<sup>f</sup> Robust regression model used due to outliers in the predictor variable. Robust regression uses estimation techniques to de-emphasize outliers; *p*-value reported from robust regression model.
*Note:* Statistically significant correlations with ASD Outcomes are denoted with bold text.

**Figure 1.**
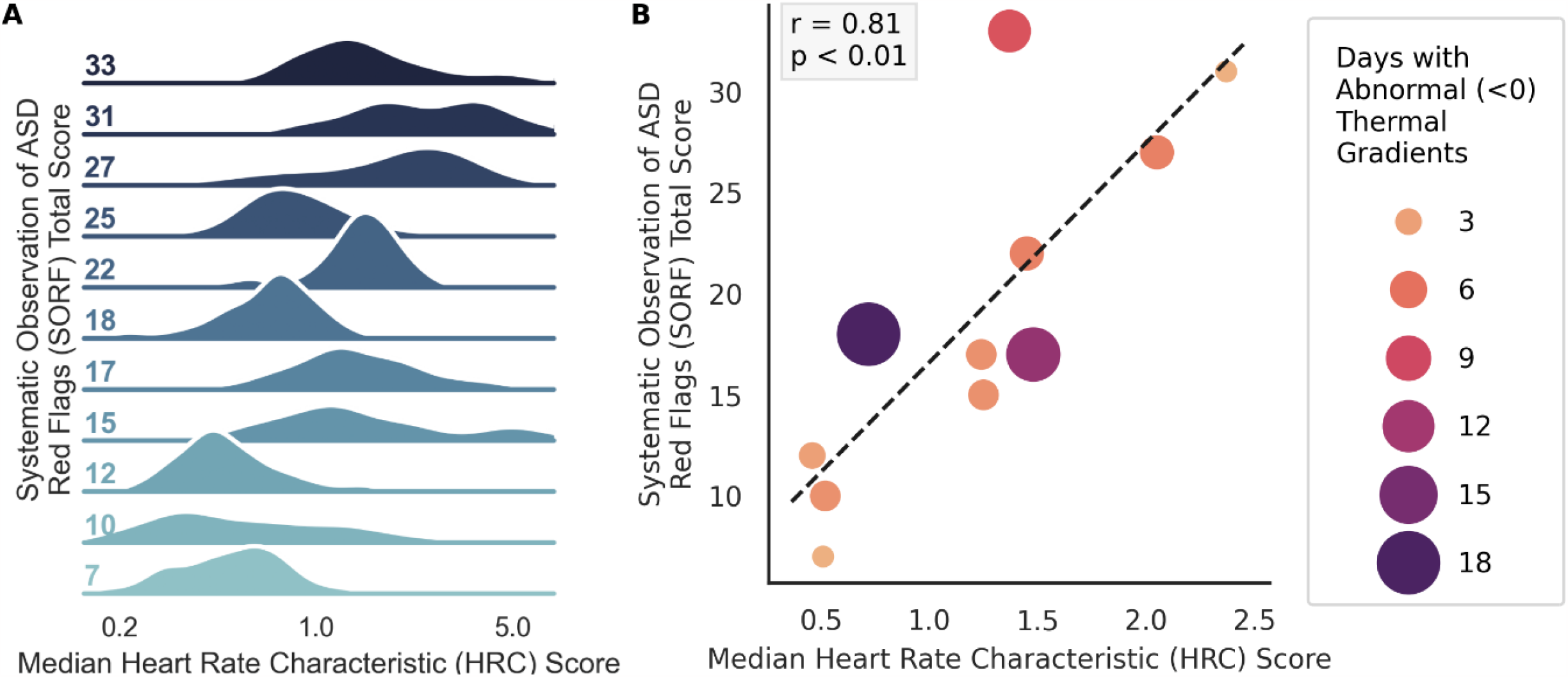
Association between neonatal abnormal heart rate characteristics (HRCs) and 1-year ASD symptoms. Higher scores indicate more HRC abnormality and more ASD symptoms. A) Ridgeline plot displaying the distribution of neonatal abnormal HRCs (HeRO scores) across participant ASD symptoms (SORF Composite score) at 1 year. B) Scatterplot and associated regression between neonatal abnormal HRCs (HeRO scores) across 1-year ASD symptoms (SORF Composite score).

## DISCUSSION

This report represents the first prospective, longitudinal study to describe predictive associations between neonatal ANS dysfunction, developmental trajectories across the first year of life, and emerging ASD symptoms at 1 year in VPT infants. The clinical presentation for most infants reflected marked delays in social communication that did not emerge until 12 months.

Significant ASD concerns were present for half of the sample by 12 months. ANS measures collected in the first month of neonatal life, more than a year prior to the ASD evaluation, were surprisingly strong predictors of ASD. Existing research points to ANS dysfunction in ASD that can be characterized by an imbalance – sympathetic overactivation and/or parasympathetic underactivation^9,10^. The strong association between ASD features and abnormal HRCs and, albeit to a lesser extent, abnormal thermal gradients, represents novel evidence contributing to an emerging theoretical model in which ANS imbalance in very early infancy may be an underlying etiological feature of ASD, particularly in VPT infants. Future studies should extend these preliminary findings by including infants born >32w GA, ANS measures that extend beyond the neonatal period, ASD outcomes at later ages, and larger sample sizes. Overall, these results highlight complementary ANS measures that describe how ANS dysfunction, likely resulting from an imbalance between the parasympathetic and sympathetic systems, may impact very early regulatory processes for neonates who later develop ASD. This finding offers a promising avenue for researching etiological mechanisms and biomarkers of ASD.

## Data Availability

All data produced in the present study are available upon reasonable request to the authors

## Availability of Data and Materials

Data is available upon request.

## Author Contributions

Drs. Jessica Bradshaw and Robin Dail conceptualized and designed the study and supervised all aspects of data collection and analysis. Drs. Jessica Bradshaw and Christian O’Reilly designed and executed the data analysis plan. Drs. Jessica Bradshaw, Robin Dail, and Christian O’Reilly drafted the initial manuscript and critically reviewed and revised the manuscript. Dr. Abbas Tavakoli contributed to data analysis and critically reviewed and revised the manuscript. Dr. Kayla Everhart, Elizabeth Dixon, and Amy Vinyard coordinated participant enrollment and retention, led data collection and data management, and critically reviewed and revised the manuscript. All authors approved the final manuscript as submitted and agree to be accountable for all aspects of the work.

## Acknowledgments

We thank the families for their time and dedication to this longitudinal research. We also thank Karen Warren for her support in hospital recruitment and Dr. John Richards for offering insight and expertise related to this work.

