## Supplementary Information for "Neonatal Autonomic Regulation as a Predictor of Autism Spectrum Disorder in Very Preterm Infants"

### METHODS

**Inclusion Criteria.** Inclusion criteria for the larger longitudinal study[1] were as follows: (a) birthweight between 500g - 1,500g, (b) born between 24-32 weeks gestational age by obstetrical dating, (c) consent and enrollment within 6 hours of birth, and (d) no immediate family history of autism spectrum disorder (ASD).

**Medical Comorbidity Score.** Medical comorbidity scores were calculated by summing the total number of conditions experienced by each infant at any time during their NICU admission, as has been done in previous studies[2]. Possible medical conditions included: infection (culture positive infection occurring < 7 days or ≥ 7 days of life), necrotizing enterocolitis (NEC) diagnosis requiring medical or surgical intervention, presence of patent ductus arteriosus (PDA) requiring medical or surgical intervention, brain injury on head ultrasound including periventricular leukomalacia and IVH grade 3 or 4, diagnosis of bronchopulmonary dysplasia or chronic lung disease (BPD/CLD), retinopathy of prematurity at stage ≥ 1 (ROP), and need for mechanical ventilation/intubation (MV).

**Abnormal Thermal Gradients.** Following study consent, two skin temperature probes (thermistors) were attached to the infant – one on their abdomen and one to the sole of one foot – with both thermistors inserted into the datalogger. Temperature data were recorded and stored every minute for the first 28 days of life, resulting in approximately 40,320 measures for each infant. Thermal gradients were calculated as the difference between the central (abdominal) and peripheral (foot) temperature (CPTd). The percentage of minutes with the foot temperature greater by any amount than the abdominal temperature was noted as a negative thermal gradient. A SAS Macro (SAS/STAT® statistical software, version 9.4.3) was used to read, combine, and analyze thermal gradients for each infant[3].

**Abnormal Heart Rate Characteristics.** Continuous cardiac activity was recorded for each infant using a standard cardiopulmonary monitor that was connected to a HeRO monitor[4]. The HeRO monitor uses continuous heart rate recording to compute heart rate characteristics that are recorded and stored. A mathematical algorithm is then used to output an hourly HeRO algorithm score indicative of abnormal heart rate characteristics (HRCs). This algorithm considers abnormality of three HRCs: standard deviation of inter-beat intervals, heart rate skewness or asymmetry, and entropy. In this study, HeRO scores greater than 1 were considered abnormal and reflect the presence of abnormal HRCs.

**Developmental Follow-Up.** Developmental follow-up assessments were administered in the child's home at adjusted ages 6, 9, and 12 months. All N=20 participants were contacted at each time point to complete developmental follow-up visits (see Fig. S1). At 6 and 9 months infants were administered the Bayley Scales of Infant and Toddler Development, 4$^{th}$ Edition[5]. The Bayly-4 includes 5 subscales represented by standard scores (mean of 10, standard deviation of 3): cognitive, fine motor, gross motor, receptive language, and expressive language. At 12 months infants were administered Mullen Scales of Early Learning (MSEL)[6] to align with a larger longitudinal study and many other studies of infants at elevated likelihood for ASD[7–9]. The MSEL includes 5 subscales represented by t-scores (mean of 50, standard deviation of 10): visual reception, fine motor, gross motor, receptive language, and expressive language. Standard scores from each measure were converted to z-scores to allow for visualization and comparison across ages and measures. The Visual Reception domain of the MSEL was used as a proxy of cognition.

**Autism Symptomology at 1 Year.** Autism spectrum disorder (ASD) is characterized by delays and differences in social communication and interaction and 2) the presence of restricted interests and repetitive behaviors[10]. Social-communication deficits are already present and measurable by 9 months of age in infants who go on to have ASD [11]. Multiple observational[12] and parent-report[13] measures are available for 1-year-old infants that quantify the presence and severity of social interaction and communication impairments and restricted interests and repetitive behaviors. These measures show good discrimination, sensitivity, and specificity for identifying ASD in at-risk infants as young as 12 months[14,15]. In alignment with existing evidence, this study measured social communication using the Communication and Symbolic Behavior Scale – Behavior Sample (CSBS-BS)[16] and measured ASD symptoms using the Systematic Observation of Red Flags for ASD (SORF)[12] and the First Year Inventory (FYI)[13].

The CSBS-BS is a 20-30-minute, semi-structured, observational assessment that is designed to provide opportunities for infants to interact and communicate with the examiner and a caregiver. It was administered in the family's home at highchair with table that was provided by the research team. The CSBS-BS was administered by trained clinicians who were trained to reliability (>80% inter-rater agreement) by the first author and by the research team of the CSBS author (A. Wetherby). The CSBS-BS provides a standardized Total Score (mean of 100, standard deviation of 15).

The SORF is an observational measure consisting of 22 items that align with DSM-5 criteria of ASD. The CSBS-BS was used as the interaction sample from which the SORF was scored. SORF scoring was completed by clinicians who were masked to participant demographic information (e.g., gestational age) and the research design, aims, and hypotheses of the current study. Clinicians were trained to reliability (>80% inter-rater agreement) by the research team of the SORF author (A. Wetherby). Each of the 22 items represents an ASD symptom or feature that is scored on a scale from 0-3, with 0 indicating the absence of the ASD feature and 3 indicating clear and definite presence of the ASD feature. A Composite Score is provided as an algorithm of selected item scores, with higher scores indicating more ASD indicators and a greater level of ASD concern. Research suggests that a cutoff Composite Score of 18 has the highest sensitivity and specificity for detection ASD in 12-month-old infants[14].

**Table S1.**

Developmental Scores at 6, 9, and 12 Months

| | 6 Months | | 9 Months | | 12 Months | |
|---|---|---|---|---|---|---|
| | Mean (SD) [a] | % Atypical[c] | Mean (SD) [a] | % Atypical[c] | Mean (SD) [b] | % Atypical[c] |
| Cognitive | 10.15 (2.54) | 15% | 7.86 (2.38) | 21% | 48.75 (11.39) | 8% |
| Gross Motor | 9.31 (2.46) | 20% | 8.00 (1.57) | 20% | 40.75 (11.99) | 50% |
| Fine Motor | 9.23 (2.17) | 15% | 9.36 (2.27) | 7% | 50.09 (15.74) | 38% |
| Receptive Language | 7.85 (1.91) | 23% | 7.14 (1.70) | 35% | 34.82 (7.76) | 64% |
| Expressive Language | 8.92 (1.38) | 0% | 8.29 (1.14) | 7% | 36.50 (12.46) | 58% |

[a] Standardized scores from the Bayley Scales of Infant and Toddler Development – 4$^{th}$ Edition. Mean of 10, standard deviation of 3.

[b] Standardized scores from the Mullen Scales of Early Learning. Mean of 50, standard deviation of 10.

[c] % Atypical represents the percent of infants at each time point who scored in the below average range.

**Figure S1.** Consort diagram of participant enrollment and subsequent visits completed at each time point. Attempts were made to contact all participants at each time point unless participant requested to withdraw from the study.

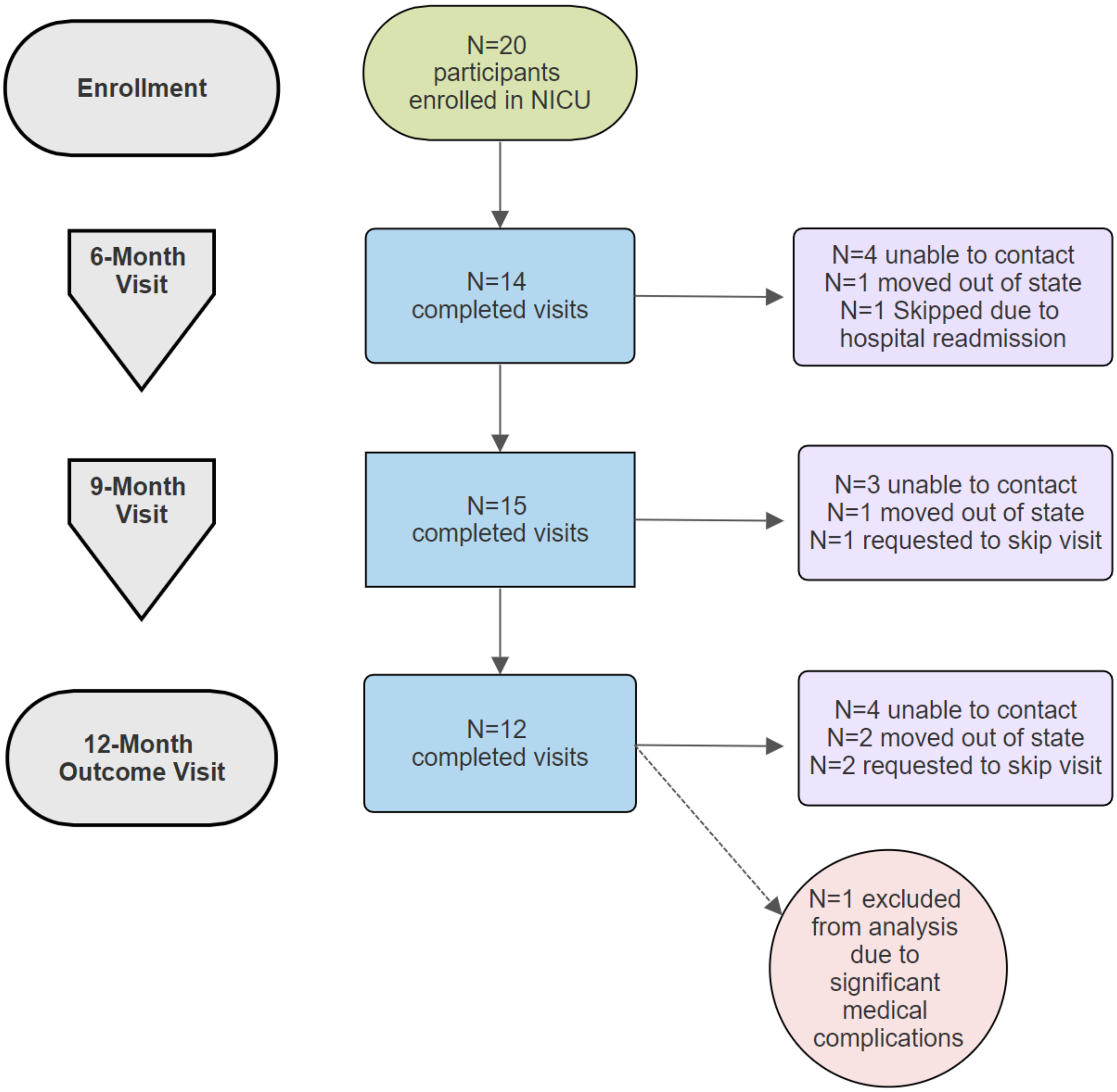

**Figure S2**. Developmental scores over time represented in z-scores where 0 is the average, -1 is one standard deviation below the mean, and -2 is two standard deviations below the mean.

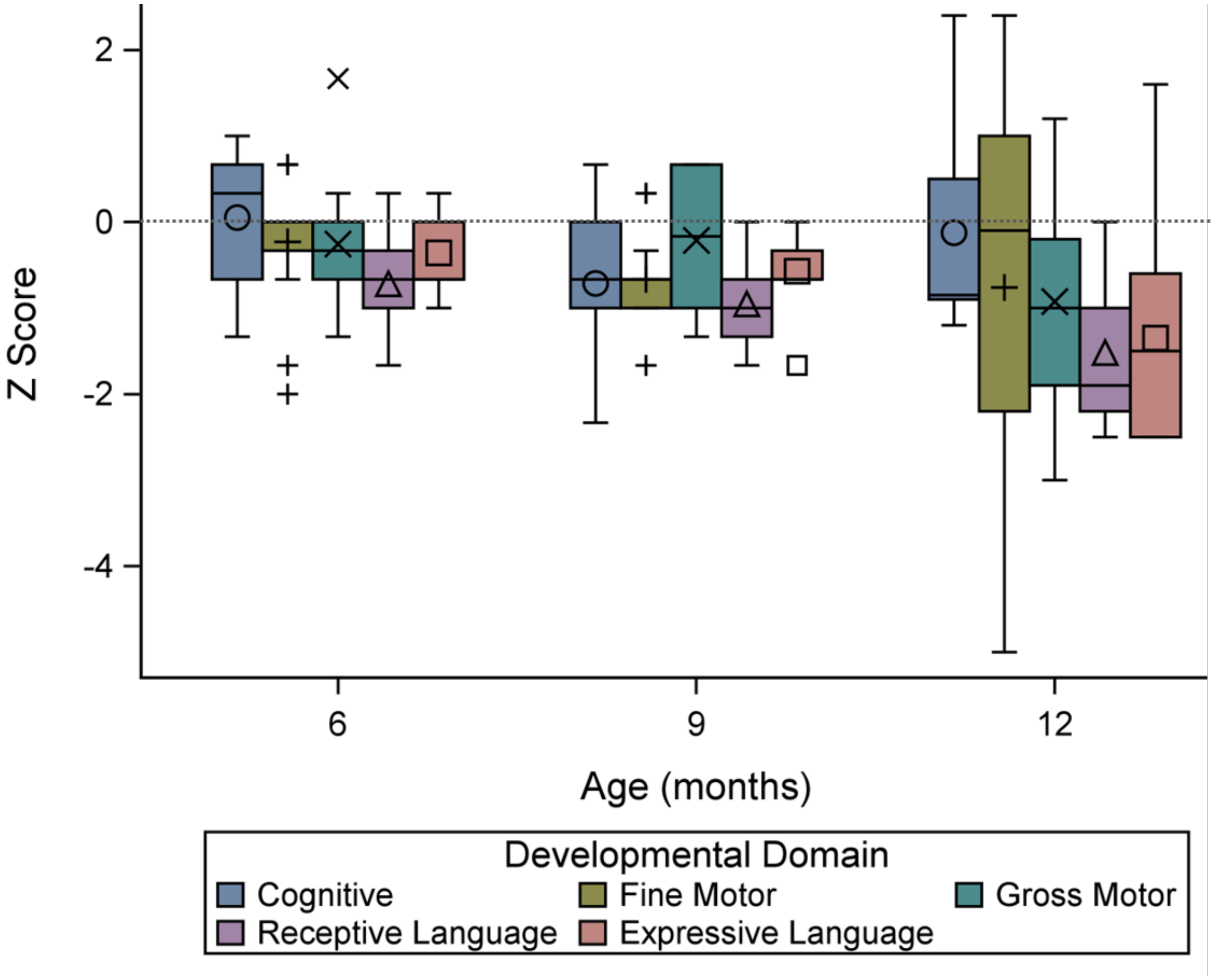
